# The Accra School Health and Environment Study (ASHES): A study of the urban environment and child health and development in Accra

**DOI:** 10.1101/2025.07.11.25331353

**Authors:** Carissa L Lange, Abosede S Alli, Kate A Kyeremateng, Jennifer F Holmes, James Nimo, Nancy Dery, Prince Anku, Lilly Adzrakor, Geofrey K Kotoka, Philomina Oppong, Jessica Kumah, Adwoa Asante-Poku, Michael Brauer, Samuel Agyei-Mensah, Sierra N Clark, Rebecca MC Spencer, Youssef Oulhote, Allison F Hughes, Majid Ezzati, Raphael E Arku

## Abstract

Elementary school and early education are crucial for children’s cognitive and social development, as well as lifetime health and well-being. For children in cities, urban schools present numerous advantages in education quality and access to resources and opportunities that stimulate learning and improve health. In Sub-Saharan African (SSA) cities, the complexity of the urban environment requires careful consideration of school environments in enhancing child health and development. Yet, little is known about environmental conditions in schools and schoolchildren’s health in rapidly urbanizing SSA cities. This paper describes the various datasets captured within the Accra School Health and Environment Study (ASHES), a study platform designed to characterize air and noise pollution at elementary schools and for schoolchildren, and their influence on key markers of childhood health and development. We outline environmental exposures and health and developmental outcomes among children living in a major metropolitan area in SSA, along with preliminary results and planned analyses.

ASHES was implemented in Accra, one of the fastest growing metropolises in SSA. Between July 2022 and May 2023, 1,037 children (∼60% female) aged 8-12 were recruited from 90 public (74%) and private primary schools. Weeklong fine particulate matter (PM_2.5_), black carbon (BC), and sound pressure levels were measured in the schoolyards. Homes of the children were geocoded and linked with spatial prediction models to estimate ambient pollutant concentrations at each child’s residence. Data were also captured on anthropometry, blood pressure, respiratory function, cognitive and behavioral functions, and sleep quality. Questionnaires gathered additional information on school, household, and sociodemographic factors. Preliminary results suggest that a third of children were hypertensive, 30% were overweight or obese, and 14% had behavioral problems. PM_2.5_ and noise levels across schools exceeded local and international standards. Several ongoing epidemiologic analyses will examine the key exposures in relation to the major outcomes.

## 1. Introduction

Sub-Saharan Africa (SSA) has the fastest growing, youngest population, and the highest urban growth rate globally [1]. The ongoing urban development and economic expansion in the region is expected to produce broad improvements in well-being and quality of life for urban dwellers [2]. For children, there is overwhelming evidence of health, development, and educational benefits from living in cities. Urban children fare significantly better in terms of mortality [3], nutrition, and development [4] compared to their rural counterparts. Even children in urban slums and informal settlements have overall health advantages over those living in rural areas. Educational outcomes in school-aged children in cities are also superior [5].

However, children in growing SSA cities are also at higher risk of exposure to urban environmental pollution in schools, a place where they spend nearly one third of their waking hours per week. Schools play a critical role in the physical, cognitive and social development of children. Yet, it is unclear how much consideration is given to air and noise pollution in urban schools in SSA and how much evidence informs the locations of schools to ensure sufficiently clean and quiet environments conducive for learning, play, and development. Schools located in high-density communities or near major roadways may have elevated average background air and noise pollution levels [6–9]. For instance, a previous study demonstrated that proximity to major roads, materials of school ground surfaces, and biomass use near schools were all determinants of students’ personal PM_2.5_ exposures [10], and another recorded mean PM_2.5_ concentrations in classrooms and schoolyards 8-fold higher than the current World Health Organization (WHO) guideline [7].

Childhood exposure to air pollution has been associated with adverse respiratory, cardiovascular, cognitive, and sleep outcomes [11–17], while exposure to noise pollution has been shown to primarily influence cognition and behavior [18,19]. Studies have also documented links between childhood exposure to air and noise pollution and reduced academic performance, with potential implications for future educational attainment and socioeconomic status [14,20–25]. The existing data underscores the need for epidemiologic evidence on the impact of multiple environmental exposures on children’s health and development in SSA’s complex urban environment. Yet, robust data on disparities in SSA’s poor urban environmental conditions and health outcomes, particularly among schoolchildren, are lacking. Such evidence will create awareness and generate policy discussions towards cleaner, quieter, healthy living environments for children in urban areas of SSA.

In Accra, Ghana’s capital and one of SSA’s fastest growing cities, recent city-wide measurements demonstrated that air and noise pollution levels vary widely and exceed local and international guidelines [26–31]. However, the impact of urban air and noise pollution and other emerging environmental exposures on markers of childhood health and development is yet to be fully appreciated in this city. This paper describes a unique research platform, designed specifically to fill this knowledge gap in the SSA context, to study both traditional (e.g. air pollution) and emerging urban environmental conditions, including noise. The Accra School Health and Environment Study (ASHES) was initiated to assess a comprehensive range of environmental exposures and markers of health and developmental outcomes in schoolchildren throughout the Greater Accra Metropolitan Area (GAMA). Overall, the ASHES project aimed to (i) measure levels of ambient PM_2.5_ and BC concentrations and environmental noise in primary schools in the GAMA, (ii) evaluate the contribution of school and residential environments to schoolchildren’s exposure to multiple air pollutants (PM_2.5_, BC, NO, and NO_2_) and noise metrics, (iii) quantify the individual and joint associations of the exposures with various markers of child health and development, and (iv) generate awareness of urban environmental health risks among stakeholders, including the school health education program of the Ghana Education Service (GES), teachers, students, and parents. The resulting data are also intended to promote science education and support policy actions that seek to achieve clean and healthy learning and living environments for children. To our knowledge, ASHES is the largest multiple air and noise pollution study to assess a wide range of childhood health markers in the urban SSA context. Additionally, ASHES can serve as a future research platform for the study of emerging environmental concerns like climate change, heat exposure, light pollution, and social networks.

## 2. Materials and Methods

### 2.1 Study location

ASHES was conducted in elementary/primary schools located within the GAMA. Covering ∼1500 km^2^ (Figure 1), the GAMA contains ∼ 6 million residents located inside multiple administrative metropolises and municipalities with the majority within the city core Accra Metropolitan Area (AMA) and the port and industrial city of Tema (TMA) to the east. Low-income/slum communities in the GAMA are densily populated with most residents living in shared compound houses and using biomass for cooking at home and for cooking food to sell on the street. Contrarily, upper-class communities consist of sparsely populated residential neighborhoods where most families live on large plots of land in modern low-rise homes and predomintalty use gas and electricity for cooking. The use of diesel generators for household and commercial activities is common in high-income neighborhoods.

**Figure 1:**
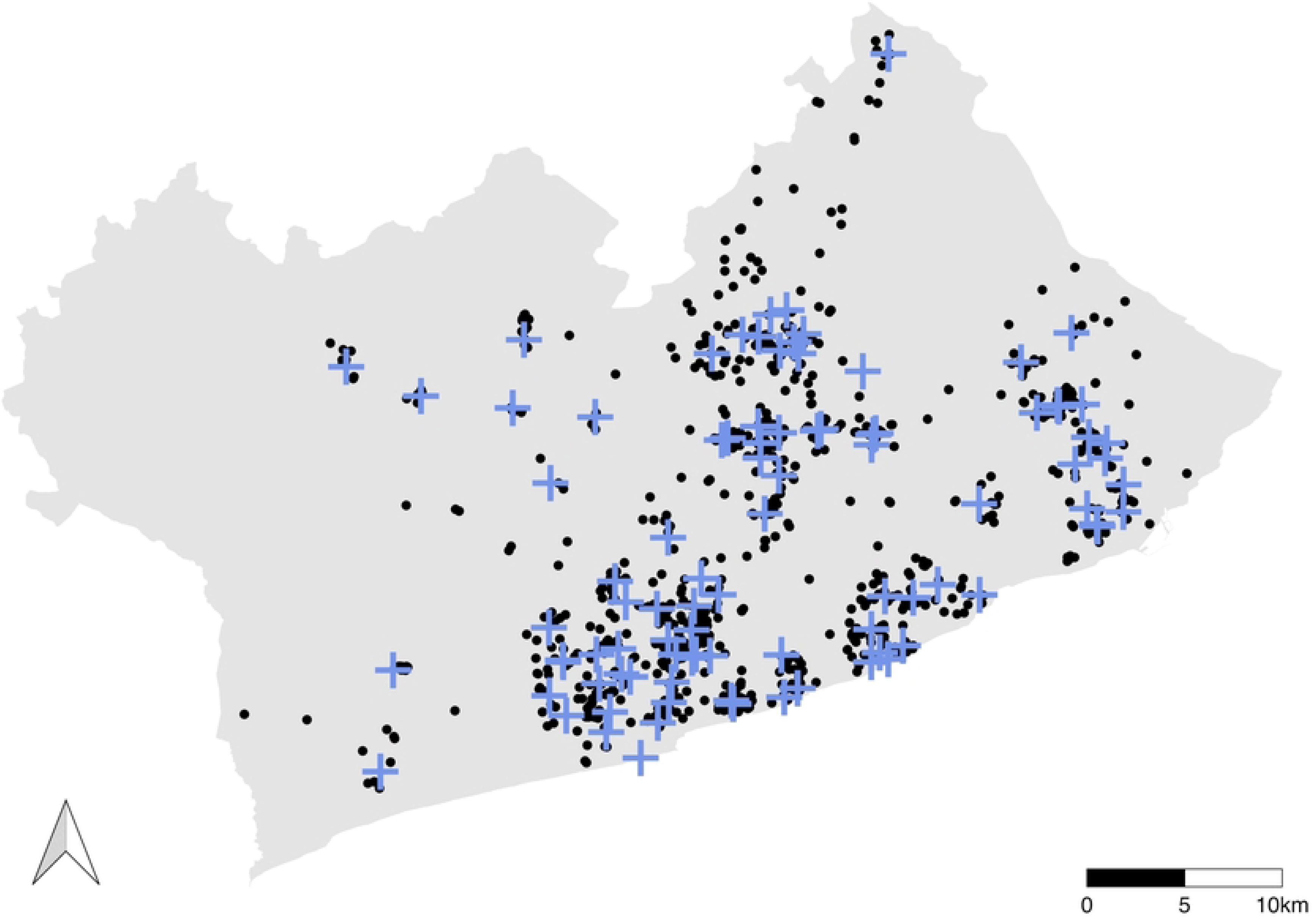
Map of GAMA and study participants’ school and home locations. In this map, GPS coordinates were displaced through a random perturbation with mean 0 and SD 0.0001 to ensure privacy of study participants.

Patterns of environmental pollution across the GAMA also follow land use features. Ambient fine particulate matter (PM_2.5_) and nitrogen dioxide (NO_2_) concentrations are 2-8-folds higher than the WHO guideline [26,27,29,30], and most residents live in areas where environmental noise levels (which are predominantly from road-traffic) exceed the WHO (European) guidelines for road-traffic noise [28,31]. In particular, residents living in the inner core of the city, commercial areas, and poorer neighborhoods are at the greatest risk of exposure [26–28]. The GAMA is also characterized by two major seasons each year: the rainy (May-September) and dry dusty Harmattan seasons, with air pollution levels substantially elevated in the Harmattan period. With average temperatures and relative humidity ranging 24 - 33° C (75-92 F) and 77-85%, respectively, heat exposure, especially night time heat is increasingly becoming a public concern. Thus, it is important to understand how these complex and considerable spatial and temporal patterns of pollution in relation to local and regional emission sources and factors influences exposures and health impacts amongst school children [32,33].

### 2.2. School system in Accra

Like the rest of the country, schools in Accra currently operate a 6-3-3-4 system, where basic education comprises of six years of elementary and three years of junior high education. This is followed by three years of senior high and the standard four-year university education. Basic education, which begins at age six and provided by both public and private schools, is compulsory for all children. Public schools are free, but they are often known to be overcrowded and lack resources. This perception facilitated the spread of private schools, particularly for upper-income earners and in areas where governmental provision is inadequate or lacking. Although more expensive, private schools are considered/perceived to offer a better quality of elementary education. The overall basic education curriculum is set centrally by GES and focuses on developing basic reading and writing abilities, and arithmetic and problem solving skills. English is the official language of instruction, although students may study in local languages for the first three years; all textbooks and materials are in English. The average school day length is eight hours. Private elementary schools generally follow GES’ curricular.

### 2.3 School selection and study population

ASHES recruited 1,037 children aged 8-12 years from 90 elementary schools across the GAMA, comprising of both public (73%) and private (27%) schools. The schools were selected from each of GAMA’s 13 metropolises and municipalities; each of these districts vary substantially by population density, land use, urbanicity, and neighborhood socio-economic status.

To select the schools, we first obtained from GES the list of all elementary schools in the GAMA and geocoded their locations using data from Google Places and handheld GPS devices, as there was no publicly available geocoded map of schools in Accra. The list included ∼700 (public) and ∼2000 (private) primary schools; we had no data on the share of enrollment in private versus public schools. Inspection of the map suggested that more elementary schools in the GAMA are located in low-income communities and localities in and around the most urbanized AMA and TMA. We used the following approach to select schools, on the basis that schools in proximity are similar in terms of neighborhood and student characteristics. For peri-urban districts, we selected four public schools per district that were not in close proximity to each other, along with one private school in each district for comparison. In the more urbanized districts within and around AMA and TMA, we selected eight or nine public schools and two or three private schools per district. This resulted in a total of 100 schools, of which ten (10%) refused participation (Figure 1). We over-represented public schools because that is where many national and local policy interventions are implemented and enforced. Although private schools generally follow GES’ curriculum, GES’s administrative policies are not necessarily followed by private school proprieters; we nonetheless included some private schools for comparison.

The schools were visited to explain the aims of the study and request participation. A total of ten 4^th^ graders from each school were specifically selected, ensuring equal representation of both genders. If this target was not met, 5^th^ graders were invited to participate to complete the sample. The students themselves were provided with assent forms detailing the expectations of the study. This age range was just old enough to accurately respond to survey questions and properly handle/maintain sensors, as fourth graders also participate in Ghana’s National Standardized Test, which assesses proficiency in numeracy and literacy in elementary schools; these outcomes are relevant for ASHES future follow-up study. Children were eligible for inclusion in the study if they had resided at their current address for at least one year, were willing to complete the children’s questionnaires, and their parents/guardians were willing to be contacted by trained research assistants and study nurses to complete the parent survey.

Locations of the participants’ homes were mapped using GPS coordinates provided by parents/guardians through a recently created and publicly available Ghana_Post_GPS (https://www.ghanapostgps.com/) system, Ghana’s official digital property addressing system. Field assistants visited the homes of those without the new Ghana_Post_GPS address and recorded their geographic coordinates using handheld GPS. The locations of the homes and schools of participants are shown in Figure 1.

### 2.4 Ethical approval

The study was approved by the Institutional Review Boards of the University of Massachusetts Amherst (Reference #: 3157) and University of Ghana, Legon (Reference #: ECH 149/18-19). Additional authorizations to conduct research within schools were obtained from different levels of the GES. We first secured approval from the National Headquarters. We then obtained additional approvals from all lower administrative levels within the GES structure, with each letter minuted to the next level, in the following order: (i) Greater Accra Regional Directorate; (ii) District and Municipal Directorate; and (iii) Heads of the participating school. The field team hand-delivered these letters and followed up with phone calls to obtain the approvals. Written informed consent was obtained from school administrators and parent/guardian(s) before taking part in this study.

### 2.5 Patient and public involvement

No patients were involved in this study. However, the study involved primary schools and schoolchildren. The research questions and outcome measures were informed by our prior fieldwork and experience in local schools and communities, that show high personal exposure among schoolchildren and heightened public concern about rising air and noise pollution and heat exposure. Consented students themselves were not directly involved in the study design or recruitment. However, the study questionnaires were piloted at two elementary schools to assess clarity and feasibility, and revisions were made based on their feedback. The teachers at the schools also provided suggestions on the implementation strategy to minimize interruption during school hours. As outlined in the consent form, summary results will be provided to schoolchildren, educators, and parents during follow-up visits.

### 2.6 Study design and data collection

A local field team consisting of two field assistants and six nurses were trained to conduct the environmental measurements and health outcomes assessments. Air and noise data were collected in schoolyards over a one-week period. For logistical reasons, monitors were deployed in five schools in each measurement week between the hours of 9 am and 11am. Measures of childhood markers of health (height, weight, blood pressure (BP), cognitive/behavioral traits, lung function) were gathered and recorded at the school, and other characteristics (sleep, physical activity, nutrition, household information, demographics, etc.) were collected from children and their parents/guardians at their residences (Supplemental Table 1).

### 2.7 Environmental assessment

#### 2.7.1 School and classroom environment survey

We gathered information on the built environment at each school (Figure 2). These included details of material on the surfaces of the schoolyard and playground (grass, paved, packed and loose dirt); classroom floor (finished, unfinished); classroom walls (finished, finished and painted, unfinished); windows in the school building (glass and openable, louvre and openable, wooden and openable, hollow blockwork); presence of separate playground (yes/no); greenness/vegetation (number of trees in the schoolyard); classroom crowding (classroom dimensions/number of children in class); and building safety (cracks in walls, floors, and windows). In addition, information was gathered on the neighborhood type in which the school resides (low/medium density, high density, commercial/business, background/other), type of road closest to the school (major, secondary, minor), the material of road surface closest to the school (paved, paved/dirt, dirt), traffic density on the nearest major road (low: 1-lane, no congestion; medium: 1-2 lanes, some congestion; and high: ≥ 2 lanes, more congestion), and commercial activity near the school. Congestion was measured at the time of instrument installation between 9-11am.

**Figure 2:**
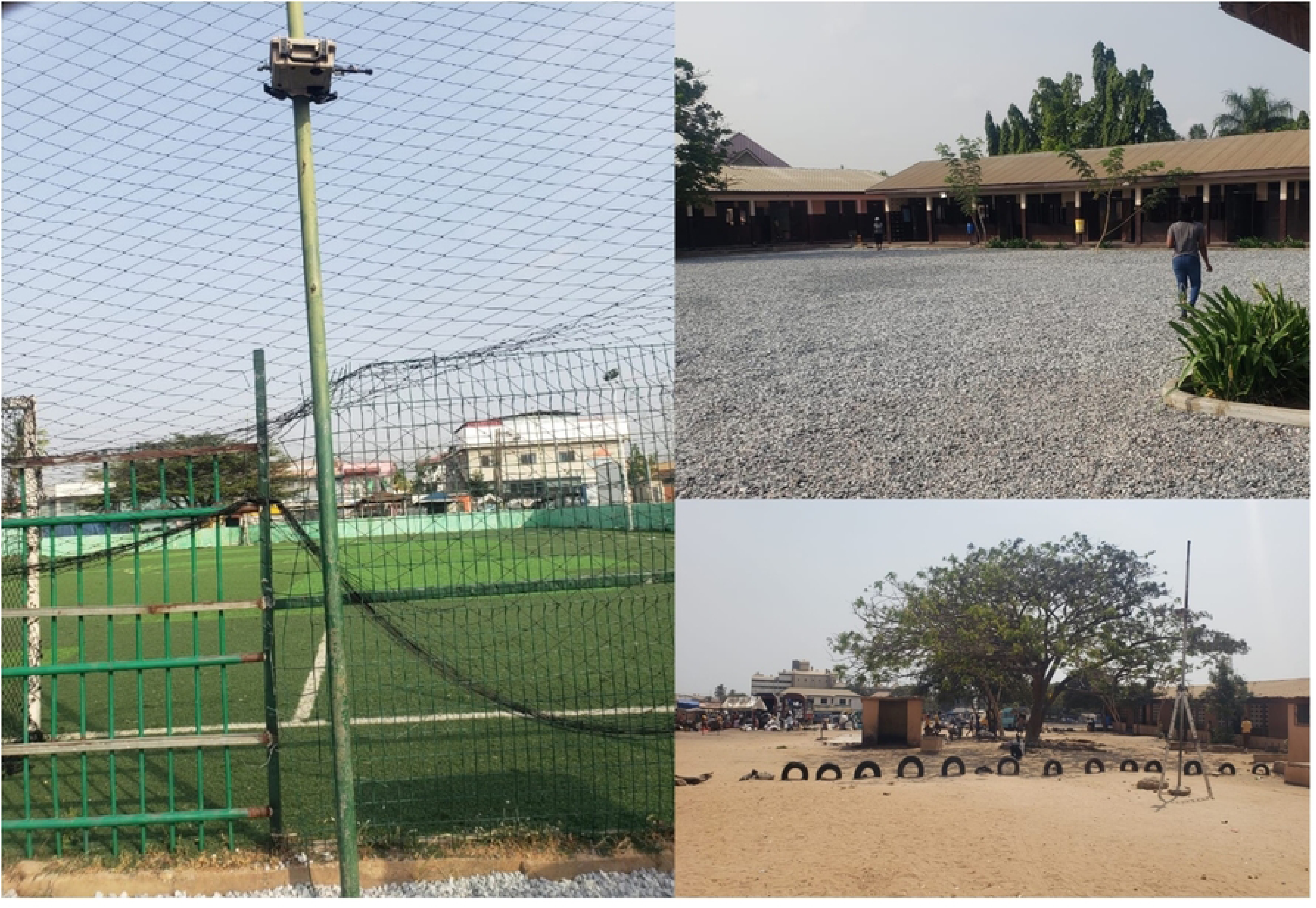
Example schoolyards in our study.

#### 2.7.2 Air and noise pollution measurements in schoolyards

We conducted week-long measurements of ambient PM_2.5_ (integrated and continuous) concentrations and sound pressure levels in the yards of the schools. Integrated (filter-based) PM_2.5_ data was collected using the Ultrasonic Personal Aerosol Sampler (UPAS) [34] while the continuous PM_2.5_ concentrations were measured using the ZeFan [35], a portable direct-reading device that is based on a light scattering technique. The light absorbance of the filters was used as proxy for BC concentration.

Previous studies in Accra have demonstrated strong seasonal variation in air pollution. Thus, data from schools monitored in different weeks throughout the study are not directly comparable. Following a previously reported approach, we adjusted each school’s weekly data for time/season by applying a temporal adjustment factor (TAF). TAF was computed separately for each measurmenet week using data from 10 outdoor monitoring sites that were continuously collecting data across the city during the study period [26,27]. Breifly, a TAF for each measurement week was calculated as the ratio of the mean PM_2.5_ or BC across all ten outdoor sites in that measurement week to the annual mean PM_2.5_ or BC across all outdoor sites. These weekly TAFs were applied to the measurements collected in schools during the corresponding week to estimate a comparable annual equivalent concentration for each school.

Outdoor sound levels were measured in one-minute integrated intervals in A-weighted decibels (dBA) using the Noise Sentry Sound Level Meter (SLM) from Convergence Instruments (Canada) [36]. Detailed information on ambient air monitors/sensors and SLMs and sampling techniques can be found elsewhere [26,28,37]. We calculated a metric of equivalent continuous sound levels from the SLM data, integrated across measurements conducted on school days (Monday – Friday) and also during typical school hours (7am – 3pm). We refer to this metric as *L_day(school)_*. Furthermore, as we wanted to characterize the environmental noise that children experienced at school, as opposed to created themselves (i.e., from laughing/playing/talking), we omit SLM data during formal school break periods (i.e. times during school hours when children go out into the school yard to play, which are typically from 9:30am – 10:00am and 12:00pm – 12:30pm). Hourly equivalent continuous sound levels were on average 0.7 dBA higher than the other school time hours.

#### 2.7.3 Estimates of air and noise pollution at participant’s residence

We could not conduct direct measurements in participants homes due to cost and duration, as is the case with most large-scale epidemiological studies [38]. However, with geo-coded information on each child’s home location, we were able to estimate annual exposures to multiple pollutants for each child by leveraging and linking to recently developed statistical predictive models for PM_2.5_, BC, NO and NO_2_ pollution [29,30], several metrics of environmental noise (LAeq_24hr_, L_den_, L_night_, L_day_) [31] and the frequency of different types of urban sounds, including road-traffic and nature-based sounds (e.g., birds) [39]. The high-resolution space-time mixed-effect models of air pollution estimated weekly concentrations at 50-meter spatial resolutions, and the environmental noise models estimated annual concentrations for different periods of the 24-hour day at the same spatial resolution. These land use regression models (LUR) were developed using city-wide yearlong measurement data collected between 2019-2020 at 146 unique locations across the GAMA, along with geospatial and meteorological variables. Details of the models, including development, validation and performance are found elsewhere [29–31]. Though the models were validated only for the period between 2019 and 2020, data was still being collected at 10 representative sites throughout this study period, which will allow us to temporally adjust the LUR models to the timing of ASHES data collection.

### 2.8 Markers of child health and development

We assessed several measures related to childhood health and development (Figure 3). These include height, weight, BP, cognitive/behavioral traits, lung function, sleep, physical activity, and dietary intake (Figure 4).

**Figure 3:**
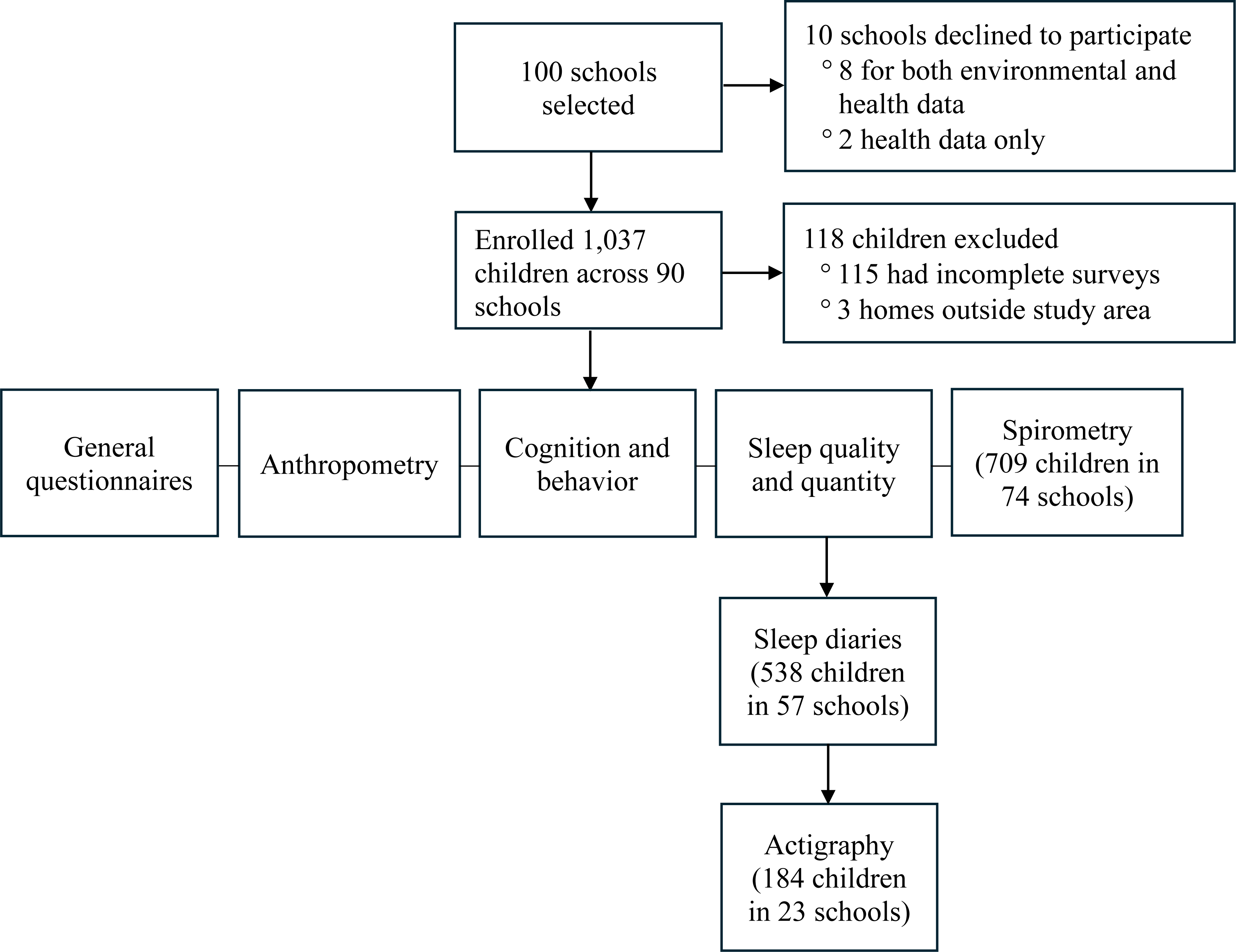
Example classroom environment and data collection. (Contact the corresponding author to request access to these images.)

**Figure 4:** Study design and number of schools and participants.

#### 2.8.1 Anthropometric measurements

Trained nurses measured each child’s height and weight, which were used to compute body mass index (BMI) as the ratio of the weight (kg) and height squared (m^2^). Height was measured to the nearest 0.1 cm while students stood barefooted on a portable stadiometer (model: Seca 216, Hamburg, Germany). Weight was recorded without shoes and bulky clothing to the nearest 0.1 kg using a calibrated digital weighing scale (model: Seca 813, Hamburg, Germany). BMI-for-age z-scores were calculated using WHO growth reference curves for children aged 5-19 years and the children were categorized as underweight, normal, overweight, or affected by obesity [40].

Systolic and diastolic BP were measured using the Omron automated BP instrument (Model: Omron HEM7131-Z, IL, USA). Children were first asked to remain in a seated position for five minutes. Afterwards, three BP measurements were taken on the left arm with a pediatric cuff at approximately 10-minute intervals [41]. Each individual record was retained but the average of all three measurements was used as the final BP value. Sex-, age-, and height-specific BP percentiles were calculated for each child using the final BP values. Following guidelines from the National Institutes of Health, BP was considered elevated if systolic and/or diastolic BP was ≥ 90^th^ percentile [42].

#### 2.8.2 Cognitive and behavioral functions

We gathered data on children’s cognitive and behavioral functions using neuropsychological assessment batteries that captured a wide range of behavioral and cognitive traits [43–45]. The Strengths and Difficulties Questionnaire (SDQ), a behavioral screening questionnaire for 2-17-year-olds, was used to capture five behavioral subscales: emotional symptoms, conduct problems, hyperactivity/inattention, peer relationship problems, and prosocial behavior. We used the parent’s version of the SDQ, which comprised of 25 items scored (range: 0 – 10) on a 3-point Likert scale: 0 (“not true”), 1 (“somewhat true”), and 2 (“certainly true”). A total difficulties score ranging from 0 to 40 was calculated by summing four of the subscales (emotional, conduct, hyperactivity, and peer). Higher total, emotional, conduct, hyperactivity, and peer SDQ scores indicate higher difficulties. Two additional subscales were calculated: internalizing problems (summed from emotional and peer subscales) and externalizing problems (summed from conduct and hyperactivity), both with scores ranging from 0 to 20.

The NIH-Toolbox Cognitive Battery (NIHTB-CB), an iPad-based battery, was used to evaluate brief memory, executive function, processing speed, and language tests. The NIHTB-CB is a computerized assessment validated in ages 3 to 85 years in the general US population and includes seven tests: Dimensional Change Card Sort (DCCS), Flanker Inhibitory Control and Attention (Flanker), List Sorting Working Memory (LSWM), Pattern Comparison Processing Speed (PCPS), Picture Sequence Memory (PSM), Picture Vocabulary (PVT), and Oral Reading Recognition (OR). The scoring, which was done automatically by the Toolbox software provided four types of composites: Fluid Cognition (scored from DCCS, Flanker, LSWM, PSM, and PCPS), Crystallized Cognition (from PVT and OR), Cognitive Function (scored from all seven tests), and Early Childhood (scored from DCCS, Flanker, PSM, and PV).

#### 2.8.3 Spirometry and respiratory health

Trained assistants conducted spirometry measurements in the children using a handheld EasyOne spirometer [46] to examine physiological lung function metrics including forced vital capacity (FVC), forced expiratory volume in 1 second (FEV1) and peak expiratory flow (PEF) [47–49]. The spirometers were calibrated at the beginning of each day, and measurements followed the current American Thoracic Society (ATS)/ European Respiratory Society (ERS) 2019 spirometry protocol [47]. In summary, participants rapidly inspired air using maximum effort. This was followed by a quick and forceful “blast” of expiration that was sustained until their lungs were completely emptied (maximum of 15 seconds). Participants then inspired at maximum flow, returning to maximum lung volume. The test was discontinued or not conducted for participants who reported being ill or having pain. Operators adhered to all hygiene protocols and participants were instructed to sanitize before and after performing the maneuver [50].

Predicted FVC, FEV1 and PEF values for participants were generated based on their demographic information data [47]. Additionally, temperature and humidity values, which were obtained from a Kestrel [51] device and the Ghana Metrological Agency website, respectively, were documented during each daily session. The EasyWare ndd Medical Technologies software provided both the study subject and the operator with a digital visualization of the flow versus volume and volume-time graphs [47,48].

#### 2.8.4 Bacteria detection on spirometry filters

Used electret filters from the spirometers were collected in efforts to determine the bacterial composition of exhaled bioaerosols on the spirometry filters. The filters were aseptically transferred from their disposable holders into sterile zip-lock bags and transported in an insulated portable cooler to the Noguchi Memorial Institute for Medical Research’s bacteriology laboratories. Samples were stored at -80 degrees prior to bacteriological processing [52].

#### 2.8.5 Sleep quality and quantity

A subset of parents/caregivers (n = 538) from 57 schools completed sleep diaries over a 2-week period for their children. To obtain the full two weeks of data, diaries were not given in schools that were visited close to exams, sports season, religious breaks, or at the end of the school term; this accounted for the reduced number of parents/caregivers and schools that completed the sleep diaries. The diaries captured information on bedtime, rise time, location of sleep (i.e., own room vs. parent’s room), sleep surface (i.e., mattress vs. mat), and number of nightly awakenings. Parents/caregivers were also asked to report any unusual events that could have influenced the previous night’s sleep (e.g., illness, medications). A smaller subset of these children (n = 172) from 23 schools wore actigraphy watches (Actiwatch Spectrum Plus, Philips Respironics, Bend, OR) to objectively measure sleep quality and quantity in terms of sleep schedule (e.g., sleep onset time, rise time), duration (e.g., time in bed, total sleep time), and fragmentation (e.g., length and number of nightly awakenings). Due to the limited number of the actigraphy watches available, schools were only invited to participate in the actigraphy substudy when the watches became available. These participants were instructed to wear the device on their non-dominant wrist continuously for the entire 2-week assessment period, resulting in 14 days and nights of actigraphy monitoring. Actigraphy is considered a reliable means of assessing sleep in children of this age [53].

#### 2.8.6 Questionnaires

For all participants, parents provided information on the presence, duration, and frequency of respiratory conditions like cough, wheeze, bronchitis, and asthma through interviewer administered surveys. Information on medication use, hospital admissions, and impact of respiratory illness on school attendance was also gathered. We additionally collected a wide range of characteristics pertaining to the home, family, parental, lifestyle, and child environments. Demographic, household, and environmental information were obtained from both schoolchildren and their parents/guardians. This included questions on maternal age and education, ethnicity, wealth, household size, cooking fuel and location, as well as child’s physical activity, diet, and perception of noise annoyance (Supplemental Table 1). The pre-tested survey questions were administered by the health team and managed using REDCap electronic data capture tools hosted at the University of Massachusetts Medical School [54–56].

### 2.9 Data handling, quality assurance/quality control

The field team received training on data capture approximately one month prior to the study recruitment and questionnaire administration. The study questionnaires were piloted at two elementary schools and revised to improve the flow and clarity of questions. Participant’s survey records were electronically captured with REDCap and checked daily for completeness, accuracy, and consistency. Signed informed consent forms were archived in a secure cabinet placed in the study laboratory, and digital copies stored in REDCap. Only authorized personnel were granted access to project REDCap page.

## 3. Preliminary results and planned analyses

Here, we present summary statistics for the exposures and health outcomes. We also provide descriptions of the ongoing epidemiologic analyses for each outcome in relation to the exposures. Each of these analyses will be published separately, and we expect a total of 8-10 manuscripts from the current work.

### 3.1 School characteristics

Ninety two schools agreed to participate in the environmental monitoring and 90 agreed to both environmental monitoring and demographic and health data collection. Of these, 26% (n = 23) were private and 74% (n = 67) were public. The average class size was 42 students but varied considerably between private (mean = 21) and public (mean= 49) schools. Similarly, public schools were more crowded than private ones (1.3 vs 2.0 m^2^ per pupil). Other variables, which captured information regarding the built environment of the schools (e.g., greenness, surface materials, neighborhood type, etc.) are presented in Supplemental Table 2.

### 3.2 Participant characteristics

Of the 1,037 children that initially provided written consent, 118 (∼11%) were excluded because (i) parents were either not reachable (due to nationwide issues with the telecommunication system in Ghana during the study period) or later declined to complete the surveys (n = 115); or (ii) residential address, which was necessary for estimating residential exposure at home, was outside the study area (n = 3). A total of 919 participants were included in ASHES for analysis. The mean (SD) age for the enrolled participants was 10.7 (1.2) years, and the majority (∼60%) were female (Table 1).

**Table 1:** General characteristics of study participants.

| Variables | All participants (N = 919) | Public (N = 691) | Private (N = 228) |
| --- | --- | --- | --- |
| Age (years) | 10.7 ± 1.2 | 10.8 ± 1.2 | 10.5 ± 1.1 |
| Male | 369 (40.1%) | 260 (37.6%) | 109 (47.8%) |
| Weight (kg) | 35.1 ± 9.0 | 34.0 ± 7.7 | 38.4 ± 11.5 |
| Height (cm) | 137.9 ± 10.9 | 135.6 ± 11.1 | 141.4 ± 9.3 |
| Body mass index (kg/m<sup>2</sup>) | 18.6 ± 3.8 | 18.5 ± 3.6 | 18.9 ± 4.3 |
| Body mass index category |  |  |  |
| *Underweight* | 30 (3.3%) | 25 (3.6%) | 5 (2.2%) |
| *Normal weight* | 611 (66.5%) | 466 (67.4%) | 145 (63.6%) |
| *Overweight* | 174 (18.9%) | 135 (19.5%) | 39 (17.1%) |
| *Obesity* | 104 (11.3%) | 65 (9.4%) | 39 (17.1%) |
| Systolic blood pressure (mmHg) | 107.2 ± 9.7 | 107.4 ± 9.5 | 106.7 ± 10.2 |
| Diastolic blood pressure (mmHg) | 69.1 ± 7.1 | 69.2 ± 7.0 | 69.1 ± 7.4 |
| Elevated blood pressure category |  |  |  |
| *Hypertension* | 163 (17.7%) | 131 (19.0%) | 32 (14.0%) |
| *Pre-hypertension* | 140 (15.2%) | 109 (15.8%) | 31 (13.6%) |
| Maternal ethnicity |  |  |  |
| *Akan* | 356 (38.8%) | 270 (39.1%) | 86 (37.7%) |
| *Ewe* | 220 (24.0%) | 146 (21.2%) | 74 (32.5%) |
| *Ga/Dangbe* | 171 (18.6%) | 147 (21.3%) | 24 (10.5%) |
| *Hausa* | 74 (8.1%) | 53 (7.7%) | 21 (9.2%) |
| *Other* | 97 (10.6%) | 74 (10.7%) | 23 (10.1%) |
| Maternal education |  |  |  |
| *Primary school or lower* | 365 (39.7%) | 324 (46.9%) | 41 (18.0%) |
| *Secondary school* | 378 (41.1%) | 280 (40.5%) | 98 (43.0%) |
| *College or University* | 176 (19.2%) | 87 (12.6%) | 89 (39.0%) |
| Cooking fuel used at home |  |  |  |
| *Clean (LPG, Electricity)* | 661 (72.0%) | 458 (66.4%) | 203 (89.4%) |
| *Biomass (Charcoal, Wood)* | 256 (27.9%) | 232 (33.6%) | 24 (10.6%) |

Data are presented as mean ± SD and number (percentage) for continuous and categorical variables, respectively.

### 3.3 Measured air pollution and environment noise in schoolyards

The median (IQR) season-adjusted measured PM_2.5_, BC, and noise levels for schoolyards are presented in Table 2. Median PM_2.5_ levels exceeded the WHO annual guideline of 5 µg/m^3^ at both private (26.0 (IQR: 22.8 - 34.1)) and public schools (30.5 (IQR: 22.2 - 39.9)), while median environmental noise levels exceeded the Ghana EPA’s standard for daytime ambient noise at educational facilities at public schools (58.0 (53.5 - 60.7)). Future analyses will characterize patterns in the measured school-level pollution by school category (public/private), location (inner-city vs suburban), and enumeration area socio economic status (low vs high) and will also evaluate potential determinants (e.g., distance to major road, schoolyard surface, vegetation/greenness). In addition, we will estimate the associations of air pollutant concentrations with the collected childhood health markers.

**Table 2:** Weeklong PM_2.5_, BC, and noise levels measured in schoolyards.

| Pollutant | All Schools | Public | Private |
| --- | --- | --- | --- |
| Air pollution |  |  |  |
| PM<sub>2.5</sub> (µg/m<sup>3</sup>) | 27.3 (22.3 - 35.8) | 26.0 (22.8 - 34.1) | 30.5 (22.2 - 39.9) |
| BC (1× 10<sup>-5</sup>m<sup>-1</sup>) | 4.3 (3.6 - 5.1) | 4.4 (3.9 - 5.3) | 3.4 (3.2 - 4.5) |
| Noise metrics |  |  |  |
| L<sub>day(school)</sub> (dBA)* | 57.2 (52.9 - 60.1) | 58.0 (53.5 - 60.7) | 54.4 (52.3 - 57.9) |
| Intermittency Ratio (%)** | 52.9 (46.2 - 58.1) | 50.8 (45.6 - 55.9) | 59.8 (50.4 - 66.0) |

Values are presented as median (IQR). PM<sub>2.5</sub> and BC concentrations have been season-adjusted.
\*L<sub>day(school)</sub>: equivalent continuous sound levels (dBA) during the day-time school hours
\*\*Intermittency Ratio (IR): IR during school time hours (%) represents how 'eventful' and 'intermittent' the sound is during this time period

### 3.4 LUR model-predicted air pollution and environment noise exposure at residence

Estimates of various ambient air pollutants (PM_2.5_, BC, NO, NO_2_) and metrics of environmental noise and frequency of different types of sounds at participants’ residences are summarized in Supplemental Table 3. Overall, median PM_2.5_ and environmental noise appear to be higher at participants’ homes than at schools. Future analyses will use these estimates to conduct some of the first epidemiological studies of the impacts of multiple environmental exposures at home on sleep outcomes, behavioral problems, cognition traits, lung and respiratory health, blood pressure, and self-reported annoyance to diverse sound sources in a SSA city. We will also examine the individual and cumulative impacts of multiple exposures and will use the combination of school measurements and estimated residential exposures to determine cumulative exposures for all participants.

### 3.5 Blood pressure

The mean (SD) systolic and diastolic BP across all the children were 107.2 (9.7) and 69.1 (7.1), respectively, with ∼33% of the children either pre- or hypertensive (Table 1). An ongoing analysis will evaluate the associations of air and noise pollution with systolic, diastolic, and elevated BP using mixed-effects models with random intercepts for school or residential area to account for potential clustering. Separate models will be developed for home PM_2.5_, BC, NO_2_ and noise metrics versus school locations. Models will be adjusted for potential confounders such as age, sex, BMI, physical activity, maternal education, age, and ethnicity, and neighborhood SES.

### 3.6 Cognitive and behavioral functions

We assessed behavioral characteristics, including total SDQ scores. Preliminary results show a median total SDQ score of 8 (IQR: 5-11), with ∼14% of the children having a score ≥ 17, which is considered above the normal threshold. Domains of cognitive function evaluated included dimensional card sort (mean=5.9; range: 0–9.6), flanker (mean = 6.4; range: 0–9.5), list sorting (mean=13.0; range: 0–26.0), pattern comparison (mean=33.8; range: 11.0–56.0), and picture sequence memory (mean = -1.13; range: -2.19–1.55). We will investigate whether air pollutants and their mixtures may impact children’s beviour and cognitive function. We will estimate the associations of school- and home-averaged air pollutant concentrations, noise metrics, separately with behavioral problem scores and cognitive outcomes, using multivariable multi-level models to account for clustering, and tested effect modification by sex.

### 3.7 Spirometry and respiratory health

With only two spirometers and slowness in orienting the children to properly perform the maneuvers, we were able to complete the test for only a subset of 709 children in 74 schools within the study duration. During the analysis, grades were assigned to test results using the acceptability criteria established by the ATS and the ERS [47]. The overall mean (SD) FEV1 and FVC were 1.6 (0.3) L and 1.9 (0.4) L, respectively; their mean ratio was 0.86 (0.08). Our planned analysis will characterize lung function metrics by school type, sex, and age. The association between ambient air pollution exposure at both the homes and the schools and lung function metrics will then be evaluated.

For the bacteria composition in the exhaled air, presumptive identification of isolated colonies will be performed by subculturing single colonies onto Uri Select Agar with overnight incubation at 37°C. Subsequently, these colonies will further subculture onto Nutrient Agar for another overnight incubation at 37°C. Bacterial colonies will then be identified using Gram staining and Matrix-assisted laser desorption/ionization mass spectrometry (MALDI-TOF MS). A unique spectrum of relative abundances of ribosomal proteins will be generated and compared to the reference from the database of the MALDI-TOF to identify organism based on score of similarity [52]. Preliminary results of 50 samples have detected the presence of Acinetobacter nosocomialis, Bacillus cereus, Bacillus pseudomycoide, Erwinia species, Escherichia coli, Franconibacter pulveris, Pseudomonoas fluorescens, Pseudomonas stutzeri, Staphylococcus aureus, Staphylococcus hominis, Streptococccus salivarius. Bacillus cereus, which commonly causes poisoning, was most prevalent, having been detected in 86% of samples. This was followed by Escherichia coli and Streptococcus salivarius, which were detected in 30% and 24% of all samples, respectively.

### 3.8 Sleep quality and quantity

Of the sub-sample of children who wore the actigraph watch, 120 had sufficient actigraphy data (defined as >7 days/nights) and had, on average, 11 nights of usable actigraphy data (mean = 11.8, SD = 1.9, range: 8-14 nights), including 8.5 (SD = 1.4) weeknights and 3.3 (SD = 0.9) weekend nights. Sleep measures will be derived from scored actigraphy records and averaged across all usable days for each participant. Ongoing analysis will estimate sleep duration, defined as the amount of time (in mins) that the child was sleeping throughout the night; sleep onset as the time that the child fell asleep; wake onset as the time of awakening in the morning; and sleep efficiency as the ratio of overnight sleep duration and the total time in bed, expressed as a percentage. In initial analysis, average nighttime sleep efficiency across participants was estimated at 88.5% (ranged: 61.4-98.0).

Planned analyses include examining the concordance between the sleep diary- and actigraphy-derived measures of sleep among the subset of participants who completed both types of datasets. Based on these results, sleep diary data may subsequently be included from those participants who did not wear actigraphy watches. Additionally, we plan to describe in more detail the sleep patterns and habits of school children in the GAMA and explore environmental, household, and health factors that may relate to those sleep measures, including air pollution, noise metrics, sleep environment (room sharing, bedsharing), household features, such as type of fuel used for cooking, and children’s respiratory health. Using additional questionnaire data, we also aim to explore the distribution of children’s chronotypes and caregivers’ knowledge and perceptions of healthy sleep practices. Lastly, we will pair the sleep data with individual cognitive and behavioral measures and neighborhood environmental measures, to see what mediating effect sleep may have between environmental pollution/noise and cognitive/behavioral outcomes.

## 4. Discussion

We described a unique research platform that captured multiple exposures and health outcomes among schoolchildren in the SSA context. ASHES dataset provides a unique and rich overview of schoolchildren’s environmental exposures and health in a growing metropolitan area in SSA. This is likely the largest study to examine environmental health among schoolchildren in West Africa, and it will be among the first to quantify the distributions of exposure to air and noise pollution in relation to spatial and socioeconomic factors in Ghana. By estimating environmental exposures at schools and homes, we are able to quantify levels of air and noise pollution experienced by schoolchildren in the city while also assessing the impacts of these exposures on a variety of markers of childhood health and developmental outcomes. The results of the ongoing analyses are expected to promote public discussion regarding how best to protect children from environmental risks in SSA uban context.

Some of the major strengths of ASHES include (i) the use of both statistical predictive models and on-site school measurements to quantify children’s exposure to air and noise pollution in the two microenvironments (home and school) where they spend the most of their time; (ii) the strategic selection of schools which ensured representation of sample size of schoolchildren from all 13 GAMA districts, featuring diverse environmental and sociodemographic profiles; (iii) the examination of multiple health endpoints including blood pressure, lung function, sleep quality, and cognitive/behavioral functions; and (iv) a high participation and survey completion rate (89%), which offers a fair glimpse of exposure and health conditions among Accra schoolchildren. ASHES data will provide information regarding the prevalence of a variety of health markers among children living in the GAMA, which can be used for future longitudinal assessments. Despite these strengths, a major limitation of ASHES is its cross-sectional design, which has the weakness of temporality between the exposures and the outcomes of interest. Still, health endpoints are generally indicative of longer-term impacts of exposures, and as such, epidemiologic analyses will utilize the newly developed LUR models to predict long-term exposures at schools and residential locations. Also, school measurements of air and noise pollution were only collected for one week during the study period in groups of five schools per week, making it difficult to compare measurements across sites. However, we have adjusted for the effect of season using data from ten fixed sites located across the city, which allowed for comparison across schools. Further, ethical considerations, we could not offer any clinical referrals or inteventional recommendations (e.g. educational material) for children with incidental findings during or follwoing our clinical assessments.

In addition to air and noise pollution, ASHES will serve as data/research platform for the study of emerging environmental concerns, such as climate change, heat exposure, light pollution, and social networks in a large metropolitan area of a major SSA city. The schools and participants will be revisited to capture climate-related risks, including temperature and heat perception, water intermittency and hydration, and other social activities like play and sports. We will also document early social environments in relation to behavior and cognition in the children, as well as sanitation and light pollution at both the school and home environement.

Overall, ASHES is timely and will support recent WHO recommendations which emphasize the need for increased monitoring data and epidemiological studies in LMICs [57]. The design and methods from ASHES can be adopted by other emerging SSA cities wishing to investigate the effects of environmental pollution on childhood health outcomes.

## Data Availability

Environmental measurement data is available through the open-source platform, Zenodo, De-identified health data can be made available to investigators with projects that fall within the overall aim of ASHES. Those interested in gaining access to the data should contact Raphael E Arku.

## Acknowledgements

The authors would like to thank all parents, children, and elementary school leaders that participated in this study and generously allowed us to install monitors in the school playgrounds. We would also like to thank the staff at the Physics Department, University of Ghana for their support in organizing the laboratory used during this project.

