## Supplemental Materials for "The Accra School Health and Environment Study (ASHES): A study of the urban environment and child health and development in Accra"

**Supplemental Table 1:** List of questionnaires and tests administered to study participants

| **Survey** | **Respondent** | **Location Completed** |
| --- | --- | --- |
| Consent Form | Parent/guardian | Home |
| Assent Form | Child | Home |
| Demographics | Parent/guardian | Home |
| Household Features | Parent/guardian | Home |
| Antenatal Care History | Parent/guardian | Home |
| Child’s Birthweight | Parent/guardian | Home |
| Food frequency | Child | School |
| Breathing/Respiratory Health | Parent/guardian | Home |
| Household Stimulation | Parent/guardian | Home |
| Anthropometry | Child | School |
| Physical Activity | Child | School |
| Noise Annoyance | Child | School |
| Sleep Behavior (SBQ) | Parent/guardian | Home |
| Knowledge, attitudes, self-efficacy, and beliefs (KASB) | Parent/guardian | Home |
| Chronotype (sleep-wake preferences) | Parent/guardian | Home |
| Strengths and Difficulties (SDQ) | Parent/guardian | Home |
| Spirometry | Child | School |
| Child Sleep Diary | Child | School/Home |
| Actigraphy watch | Child | School/Home |

**Supplemental Table 2:** Environmental conditions of school sites

| **Variables** | **All schools (N = 92)** | **Public (N = 67)** | **Private (N = 23)** |
| --- | --- | --- | --- |
| Class size (# of students) | 42 (6-114) | 49 (22-114) | 21 (6-40) |
| Classroom size (m^2^) | 49 (14-78) | 54 (32-77) | 36 (14-78) |
| *m^2^ per student* | 1.5 (0.5-5.2) | 1.3 (0.5-2.8) | 2.0 (0.8-5.2) |
| Time spent in school (hours) | 7.1 ± 0.9 | 7.1 ± 0.9 | 7.0 ± 0.6 |
| Schoolyard surface material |  |  |  |
| *Packed/loose dirt* | 43 (46.7%) | 41 (61.2%) | 2 (8.7%) |
| *Paved* | 40 (43.5%) | 19 (28.4%) | 19 (82.6%) |
| *Tiled* | 2 (2.2%) | 0 (0%) | 2 (8.7%) |
| *Grass** | 7 (7.6%) | 7 (10.4%) | 0 (0%) |
| Playground surface material |  |  |  |
| *Packed/loose dirt* | 49 (53.3%) | 47 (70.2%) | 2 (8.7%) |
| *Paved* | 30 (32.6%) | 10 (14.9%) | 18 (78.3%) |
| *Tiled* | 2 (2.2%) | 0 (0%) | 2 (8.7%) |
| *Grass* | 11 (11.9%) | 10 (14.9%) | 1 (4.3%) |
| Greenness****** |  |  |  |
| *None* | 33 (35.9%) | 15 (22.4%) | 17 (74.0%) |
| *Low* | 34 (57.6%) | 30 (44.8%) | 3 (13.0%) |
| *Medium* | 18 (30.5%) | 15 (22.4%) | 3 (13.0%) |
| *High* | 7 (11.9%) | 7 (10.4%) | 0 (0%) |
| Classroom wall material |  |  |  |
| *Finished - unpainted* | 27 (29.3%) | 13 (19.4%) | 13 (56.5%) |
| *Finished - painted* | 60 (65.2%) | 52 (77.6%) | 7 (30.4%) |
| *Unfinished* | 5 (5.5%) | 2 (3.0%) | 3 (13.1%) |
| Window material |  |  |  |
| *Glass and openable* | 4 (4.3%) | 0 (0%) | 4 (17.4%) |
| *Louvre and openable* | 28 (30.4%) | 15 (22.4%) | 11 (47.8%) |
| *Wooden and openable* | 12 (13.1%) | 12 (17.9%) | 0 (0%) |
| *Hollow blockwork* | 34 (37.0%) | 33 (49.3%) | 1 (4.4%) |
| *Other* | 14 (15.2%) | 7 (10.4%) | 7 (30.4%) |
| Neighborhood type |  |  |  |
| *Commercial* | 14 (15.2%) | 12 (17.9%) | 2 (8.7%) |
| *High-density* | 15 (16.3%) | 14 (22.4%) | 0 (0%) |
| *Med/low-density* | 63 (68.5%) | 40 (59.7%) | 21 (91.3%) |
| Traffic volume near school |  |  |  |
| *High* | 8 (8.7%) | 8 (11.9%) | 0 (0%) |
| *Medium* | 17 (18.5%) | 14 (20.9%) | 3 (13.0%) |
| *Low* | 67 (72.8%) | 45 (67.2%) | 20 (87.0%) |
| Nearest road type |  |  |  |
| *Major* | 4 (4.4%) | 4 (6.0%) | 0 (0%) |
| *Secondary* | 30 (32.6%) | 27 (40.3%) | 3 (13.0%) |
| *Minor* | 58 (63.0%) | 36 (53.7%) | 20 (87.0%) |
| Nearest road surface |  |  |  |
| *Dirt* | 17 (18.5%) | 12 (17.9%) | 5 (21.7%) |
| *Paved* | 66 (71.7%) | 52 (77.6%) | 12 (52.2%) |
| *Dirt/paved* | 9 (9.8%) | 3 (4.5%) | 6 (26.1%) |

Data are presented as mean (range) and number (percentage) for continuous and categorical variables, respectively.

*Includes any presence of grass (grass/loose dirt, grass/paved, and grass)

**Greenness was defined by the number of trees present (low: 1-10, medium: 11-20, high: 21-31)

**Supplemental Table 3:** Land Use Regression (LUR) Model predicted home air pollution concentrations and noise metrics*

| Pollutant | Median (IQR) |
| --- | --- |
| Air pollution |  |
| *PM_2.5_ (µg/m^3^)* | 33.8 (32.0 - 35.4) |
| *BC (1× 10^-5^m^-1^)* | 5.2 (4.6 - 6.0) |
| *NO_2_ (µg/m^3^)* | 55.5 (45.1 - 64.1) |
| Noise metrics |  |
| *L_den_ (dBA)* | 63.9 (61.0 - 67.0) |
| *L_day_ (dBA)* | 62.6 (59.6 - 65.8) |
| *L_night_ (dBA)* | 54.2 (51.6 - 57.2) |
| Sound sources |  |
| Road-traffic sounds during day-time (%) | 74.5 (65.4 - 79.3) |
| Road-traffic sounds during night-time (%) | 53.4 (40.1 - 61.3) |
| Animal (nature-based) sounds during day-time (%) | 32.5 (20.9 - 46.8) |
| Animal (nature-based) sounds during night-time (%) | 50.2 (39.9 - 62.2) |

*****LUR models were developed using city-wide yearlong measurement data (2019-2020) at 146 sites. Model predictions were linked using geo-coded locations of home residences.
